# Epigenome-wide association study using peripheral blood leukocytes identifies genomic regions associated with periodontal disease and edentulism in the Atherosclerosis Risk in Communities Study

**DOI:** 10.1101/2023.02.09.23285711

**Authors:** Naisi Zhao, Flavia Teles, Jiayun Lu, Devin C. Koestler, James Beck, Eric Boerwinkle, Jan Bressler, Karl T. Kelsey, Elizabeth A. Platz, Dominique S. Michaud

**Affiliations:** Department of Public Health & Community Medicine, Tufts University School of Medicine, Tufts University, Boston, MA; Department of Basic & Translational Sciences, University of Pennsylvania, Philadelphia, PA; Department of Epidemiology, Johns Hopkins Bloomberg School of Public Health, Baltimore, MD; The Sidney Kimmel Comprehensive Cancer Center at Johns Hopkins, Baltimore, MD; Department of Biostatistics & Data Science, University of Kansas Medical Center, Kansas City, KS; University of Kansas Cancer Center, Kansas City, KS; Division of Comprehensive Oral Health/ Periodontology, Adams School of Dentistry, University of North Carolina, Chapel Hill, NC; Human Genetics Center, School of Public Health, University of Texas Health Science Center at Houston, Houston, TX; Human Genome Sequencing Center, Baylor College of Medicine, Houston, TX; Department of Epidemiology, Brown University, Providence, RI; Department of Pathology and Laboratory Medicine, Brown University, Providence, RI

**Keywords:** Periodontitis, periodontal disease, DNA methylation, Illumina Infinium HumanMethylation450K, epigenome-wide association study (EWAS), peripheral blood

## Abstract

**Aim:** Our goal was to investigate individual susceptibility to periodontitis by conducting an epigenome-wide association study using peripheral blood.

**Materials and Methods:** For this analysis, we included 1077 African American and 457 European American participants of the Atherosclerosis Risk in Communities (ARIC) study who had completed a dental examination or reported being edentulous at visit 4 and had available data on DNA methylation. DNA methylation levels were compared by periodontal disease severity and edentulism to identify differentially methylated regions (DMRs) and evaluate the CpGs belonging to those DMRs using multinominal logistic regression.

**Results:** We identified a region in gene *ZFP57* (6p22.1) that was significantly hypomethylated in severe periodontal disease compared to no/mild periodontal disease in European American participants. A separate region in an unknown gene (located in Chr10: 743,992-744,958) demonstrated significant positive association with edentulism compared to no/mild periodontal disease in African American participants. Four CpGs in a region located within *HOXA4* were significantly hypermethylated in severe periodontal disease compared to no/mild periodontal disease in African American participants.

**Conclusions:** Our study highlights epigenetic variations in *ZPF57* and *HOXA4* that were significantly and reproducibly associated with periodontitis. Future studies should evaluate gene regulatory mechanisms in the candidate regions of these loci.

**CLINICAL RELEVANCE:** 

**Scientific Rationale for Study:** Without altering the DNA sequence, epigenetic effects (e.g., DNA methylation changes) can alter gene activity and influence host response to periodontal infections. Our well-powered study investigates individual susceptibility to periodontitis by conducting a thorough assessment of periodontitis-related DNA methylation levels in blood.

**Principal Findings:** We identified two gene regions, *ZPF57* and *HOXA4*, that are differentially methylated in individuals with compared to those without periodontitis.

**Practical implications:** Studying differential leukocyte DNA methylation patterns may point to candidate regions and underlying gene regulatory mechanisms that play a key role in the progression and/or susceptibility to periodontitis.

## Introduction

Over 47% of US adults have some form of periodontitis, and the incidence of periodontitis is rising steeply in adults 50 and older.^1-3^ Given the public health impact of periodontal disease^4,5^, studying individuals’ susceptibility to periodontitis could be crucial to building effective disease prevention strategies. In the last two decades, genome-wide association studies have attempted to discover genes that contribute to the pathogenesis of periodontitis; but these studies have found no common causal allele that reached genome-wide significance.^6,7^ Epigenetic modifications of DNA can affect the genetic blueprint of host responses, being either directly lesion-associated in target tissue or associated with an altered immune response; thus, it is of particular interest to assess epigenetic alterations in whole blood as it may (directly or indirectly) modulate the development and progression of periodontitis.

In the context of periodontal disease, biological mechanisms leading to periodontitis are driven by infections,^8^ and the host response plays a critical role in individual response. Given that changes in methylation levels can reveal a particular host response, studies of DNA methylation may yield valuable information about disease susceptibility to periodontitis. Two epigenome-wide studies (EWAS) of peripheral blood leukocytes in periodontitis have been published. A cross-sectional study in adult female twins by Kurushima et al.^9^ identified several CpGs, including one in *ZNF804A*, associated with gingival bleeding and tooth mobility. A pilot study by Hernandez et al.^10^, conducted in 8 periodontitis patients and 8 matched controls, reported statistically significant differences in peripheral blood DNA methylation in multiple regions, including the *ZNF718, HOXA4*, and *ZFP57* genes.

To expand on previous work of periodontitis-related DNA methylation changes, we conducted an EWAS to discover and validate aberrant patterns of blood-derived DNA methylation in periodontal disease severity and edentulism among 1534 Black and White older adults who underwent a clinical dental examination as a component of the community-based ARIC study. Our study aimed to identify region- and CpG-specific differential methylation in peripheral blood leukocytes collected from participants with moderate or severe periodontitis compared to those with no or no/mild periodontitis, controlling for relevant health conditions, socioeconomic status, self-reported smoking, and methylation-predicted packyears.

## Material and Methods

### Study Population

Study participants were enrolled in the Atherosclerosis Risk in Communities (ARIC) Study (RRID: SCR_021769), a prospective cohort study of cardiovascular disease risk that enrolled 15,792 participants between 1987 and 1989 from four US communities (Jackson, MS; Washington County, MD; suburban Minneapolis, MN; and Forsyth County, NC).^11,12^ Participants underwent a baseline clinical examination (Visit 1) and subsequent follow-up clinical exams (Visits 2–9 completed & Visit 10 planned). Participants provided written informed consent at each visit. The ARIC Study protocol was approved by institutional review boards at the four field centers and coordinating center.

At Visit 4 (1996–1998), participants who had at least one tooth or dental implant and were otherwise eligible and consented underwent a clinical dental examination by trained personnel (n = 6017). Edentulous participants were not eligible for the dental examination. Our analysis focused on a sample of ARIC participants with available data on leukocyte DNA methylation levels collected at Visit 2 (1990-92) or Visit 3 (1993-1995). Among the participants with methylation data, those who attended a dental examination^13^ (n = 1174) or who self-reported being edentulous (n = 360) during Visit 4 were eligible for our study.

### DNA Methylation Assessing and Preprocessing

The procedures for DNA methylation assessment have been previously described.^14^ Briefly, genomic DNA was extracted from peripheral blood leukocyte samples using the Gentra Puregene Blood Kit (Qiagen) and bisulphite conversion of 1 ug genomic DNA was performed using the EZ-96 DNA Methylation Kit (Deep Well Format) (Zymo Research).^14^ Details are summarized in the Supplemental Methods File.

### Periodontal Measures

Periodontal severity was determined based on standardized dental measurements. Probing depth and gingival recession were measured at six sites on all teeth present. Clinical attachment loss (AL) was calculated based on these two measures.^15,16^ We acknowledged the complexity of classifying periodontal disease by utilizing two different measures of periodontitis severity. The first definition, “periodontitis (ARIC),” was previously used in ARIC to categorize individuals based on the percentage of sites with AL>= 3 mm into no/mild, moderate, or severe periodontitis.^17^ The second measure, “periodontitis (CDC-AAP),” was developed for population-based surveillance of periodontitis by the US Centers for Disease Control and Prevention–American Academy of Periodontology (CDC-AAP). It includes AL and pocket depth (PD) measurements and the site location (interproximal) to classify participants into no, mild, moderate, or severe periodontitis.^18^ We used edentulism as a surrogate for the most severe type of periodontitis. ARIC participants who self-reported as both not having any natural teeth and not having any dental implants were classified as edentulous for this analysis.

### Covariate Assessment

Modifiable risk factors associated with periodontitis and edentulism include: tobacco smoking^19^, nutrition^20^, psychological stress and depression^21^, obesity, diabetes^22^, and metabolic syndrome.^23^ Data on cigarette smoking status (current, former, never) and cigarette smoking dose was measured at each visit during follow-up. Diabetes at baseline was defined as fasting plasma glucose ≥126□mg/dl, non-fasting plasma glucose ≥200□mg/dl, receiving diabetes treatment, or self-report of a diabetes diagnosis by a physician. A lifetime socioeconomic status (SES) variable, calculated based on 12 factors associated with SES, was used to identify participants with high life-course SES.^24^ Leukocyte cell composition was measured using a DNA methylation deconvolution procedure (see Supplemental Methods), given its potential to confound analyses of blood-based DNA methylation data. Lastly, we used the R function EstDimRMT to determine the number of principal components with non-zero eigenvalues needed to correct batch effects in the current analysis.^25,26^

### Statistical Analysis

We examined the association between CpG-specific DNA methylation and periodontal disease by conducting an epigenome-wide association analysis (EWAS) using multivariable logistic regression models. Such models were used to estimate odds ratios (OR) of severe vs. no/mild periodontitis (ARIC definition), moderate vs. no/mild periodontitis (ARIC definition), and edentulous vs. no/mild periodontitis (ARIC definition), or severe vs. no periodontitis (CDC definition), mild/moderate vs. no periodontitis (CDC definition), and edentulous vs. no periodontitis (CDC definition) per 1 SD increase in methylation level at single CpG sites. All models were adjusted for age, sex, five surrogate variables for batch effects, smoking category (never, former, current), packyears, BMI, and leukocyte cell composition. In addition, to control for potential residual confounding by smoking, we included a methylation-predicted packyears developed based on a meta-analysis of association results between DNA methylation and cigarette smoking in individuals from 16 cohorts.^27^ The Benjamini-Hochberg method was used to calculate the false discovery rate (FDR) for multiple comparison adjustments (FDR q-value < 0.05). Lastly, we conducted EWAS separately for African American and European American participants. Since blood samples from African American and European American participants were assayed in separate Illumina runs and at different times, we treated methylation data of African American and European American participants as two separate cohorts.

Using the DMRcate R/Bioconductor package, we conducted differentially methylated region (DMR) analyses to identify DMRs associated with periodontal disease status. A linear model that adjusted for the same covariates as the single CpG analyses was fit using the DMRcate function along with these parameters: lambda = 1,000 and kernel adjustment C = 2.^28^ Statistically significant DMRs were required to have a minimum of two statistically significant single CpGs and to meet the multiple testing adjustment criteria of FDR (q-value) <0.05 (calculated using the Benjamini-Hochberg method).^29^

CpGs belonging to the statistically significant DMRs were evaluated using a baseline category logit model using both the CDC-AAP and the ARIC periodontitis definitions. In a prior study, Hernandez et al. conducted a pilot EWAS in 8 periodontitis patients and 8 matched controls and reported statistically significant differences in peripheral blood DNA methylation in multiple regions, including the *ZNF718, HOXA4*, and *ZFP57* genes.^10^ These three DMRs were the only regions that were found to be significant with multiple consecutive CpGs in the same direction after controlling for cell composition in the model.^10^ We compiled CpGs from the three DMRs identified by Hernandez et al. and tested them in the ARIC dataset. All models controlled for age, sex, SES, BMI, diabetes status, smoking category (never, former, current), packyears, methylation-predicted packyears smoked, leukocyte cell composition, ARIC field center, and five surrogate variables for batch effects. Analyses were conducted separately for African American and European American participants.

## Results

### Population Characteristics

Table 1 summarizes the characteristics of ARIC participants included in this study by periodontal disease status defined by the CDC-AAP and by the ARIC periodontitis definitions. The average time between blood drawn and Visit 4 was 6 years. Participants with severe periodontitis were more likely to be male, black, obese, less educated, current smoker, and have lower life-course SES (Table 1). Compared to those of European ancestry, African American participants in this study were more likely to be diabetic, have low SES, are less educated, have fewer teeth, and are more likely to have severe periodontitis (Supplemental Table 1). African American participants were younger, with a median age of 55, compared to 58 years old for European American participants. 28.6% of African American participants had severe periodontal disease, with 45.7% having no/mild periodontal disease, while 11.4% and 60.2% of European American participants had severe or no/mild periodontal disease, respectively.

**Table 1.**
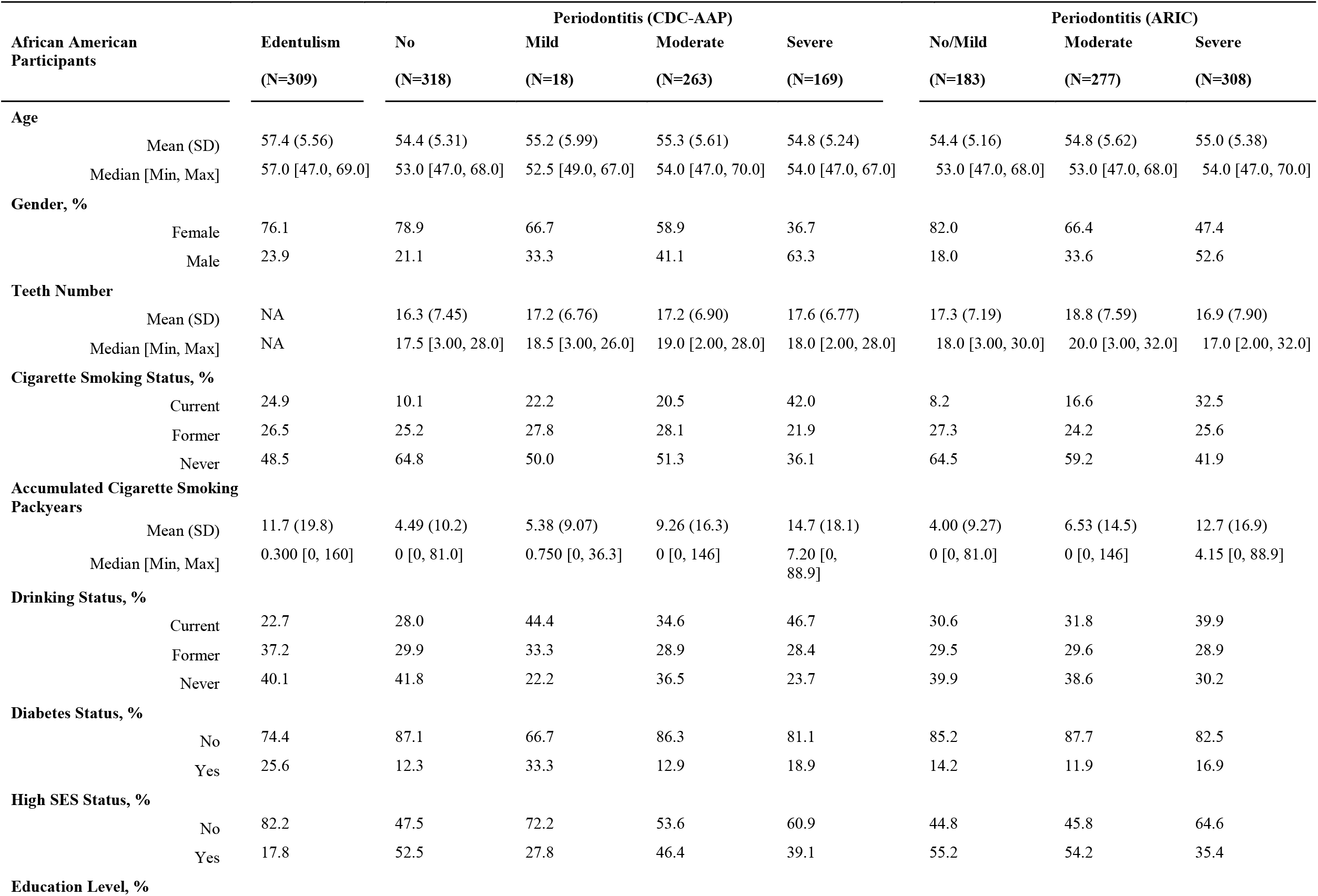
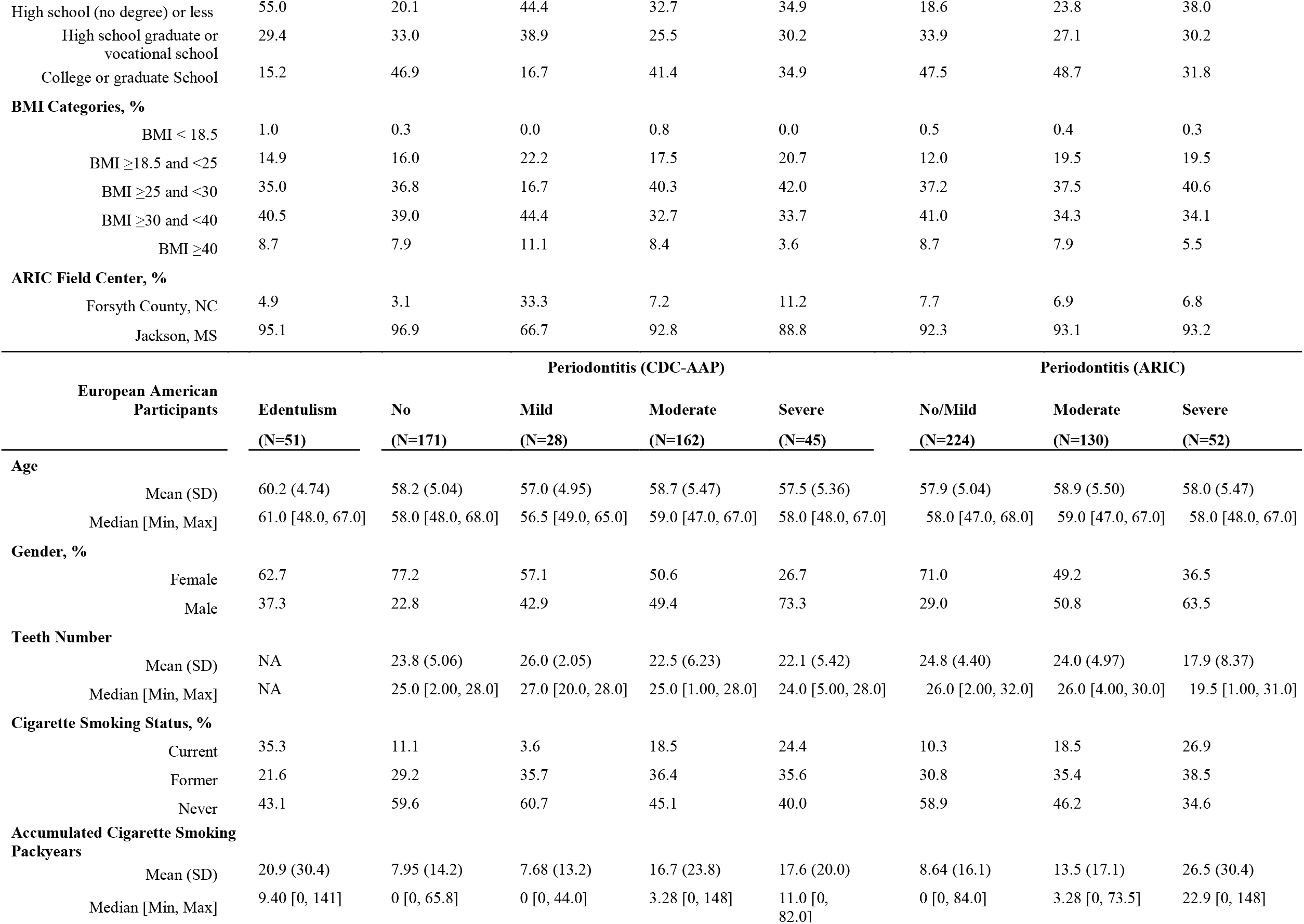
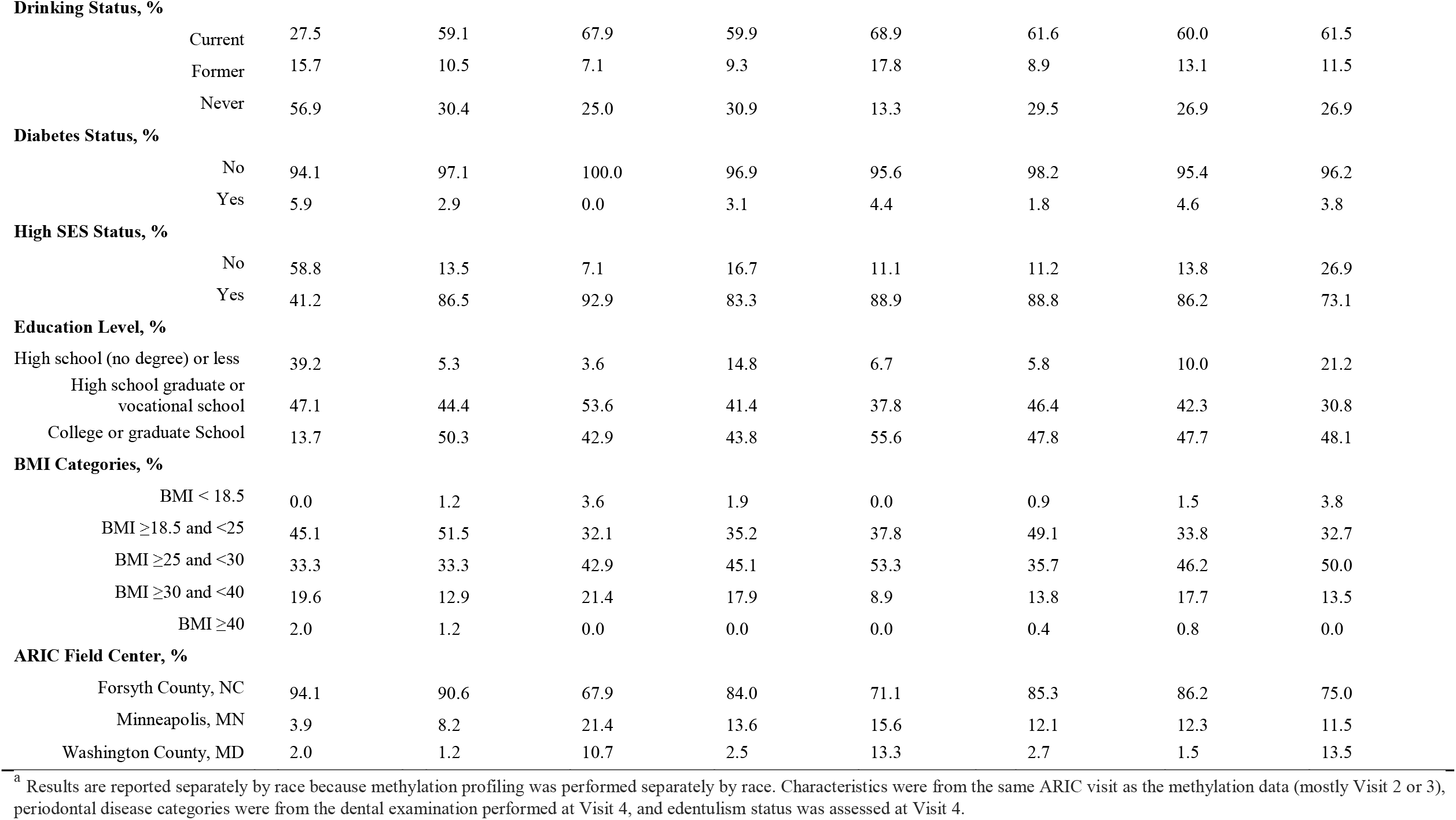
Characteristics across periodontal disease categories among ARIC participants, separately by race ^a^.

### EWAS & DMRcate Analysis

Using the ARIC periodontitis definition, the single CpG EWAS did not identify differentially methylated CpGs that were statistically significant after multiple comparison corrections (q < 0.05). After adjusting for methylation-predicted packyears, we identified one statistically significant DMR (*ENSG00000231601* manual annotation Chr10: 743,992-744,958) in edentulism when compared to no/mild periodontal disease in African American participants and one statistically significant DMR (*ZFP57* 6p22.1) in severe periodontal disease compared to no/mild periodontal disease in European American participants (Supplemental Table 2). The EWAS and DMRcate results did not vary from those presented above when using the CDC-AAP periodontitis definition for analysis.

#### Zinc Finger Protein Gene 57 (ZFP57)

The statistically significant DMR located on Chr6 (*ZFP57* gene), consisting of 17 CpGs, differed between European American participants with severe periodontal disease and those with no/mild periodontal disease. The 17 CpGs perfectly overlapped with a 21-CpG *ZFP57* region which was reported by Hernandez et al. as hypomethylated when comparing periodontitis patients with age-matched and sex-matched periodontally healthy controls living in the Republic of Colombia.^10^ Overlapping CpGs located in the *ZFP57* DMR region were found to be associated with a lower risk of severe periodontal disease compared to no/mild periodontal disease (ARIC definition) in European American participants (Table 2). This association was stronger in European American participants 58 years or older (Table 3). It was also comparable for male and female participants and participants of higher or lower SES (Supplemental Table 3). In addition, there was a statistically significant decreased risk of moderate vs. no/mild periodontal disease among European American participants 58 years or older (Table 3). The *ZFP57* CpGs were not significantly associated with risk of moderate or severe periodontal disease, as compared to no/mild periodontal disease in African American participants. However, increasing methylation of three *ZFP57* CpGs (cg11383134, cg15570656, and cg13835168) was associated with a significant decreased risk of severe vs. no/mild periodontal disease in African American participants 58 years or older (Table 3). When we used the CDC-AAP definition to categorize periodontal disease status, increasing methylation of the *ZFP57* CpGs was found to be associated with a significant decreased risk of mild/moderate vs. no periodontal disease in European American ARIC participants. The pattern of associations for those 58 years and older, male and female participants, participants of higher or lower SES, and for edentulism were all similar to what we found using the ARIC definition (Table 2 and Supplemental Table 3).

**Table 2.** Odds ratio for CpGs in the *ZFP57* ^a^ differentially methylated region and using two definitions of periodontal disease severity in ARIC ^b^.

| CpG | Odds Ratio per 1 SD increase in methylation level (95% CI) |  |  |  |  |  |
| --- | --- | --- | --- | --- | --- | --- |
|  | Periodontitis (CDC-AAP definition)<sup>c</sup> |  |  | Periodontitis (ARIC definition)<sup>c</sup> |  |  |
|  | mild/moderate vs. no | severe vs. no | edentulism vs. no | moderate vs. no/mild | severe vs. no/mild | edentulism vs. no/mild |
| **EA Participants (Visit 02)** |  |  |  |  |  |  |
| cg07134666 | **0.69 (0.54, 0.88)** | 1.09 (0.72, 1.64) | 1.00 (0.66, 1.50) | 0.84 (0.66, 1.07) | **0.66 (0.47, 0.92)** | 1.05 (0.71, 1.55) |
| cg11383134 | **0.65 (0.50, 0.84)** | 1.04 (0.66, 1.65) | 1.06 (0.66, 1.72) | **0.76 (0.60, 0.98)** | **0.65 (0.46, 0.91)** | 1.12 (0.71, 1.78) |
| cg22494932 | **0.62 (0.47, 0.82)** | 1.08 (0.65, 1.79) | 1.04 (0.60, 1.79) | **0.74 (0.58, 0.94)** | **0.69 (0.50, 0.96)** | 1.14 (0.68, 1.90) |
| cg15570656 | **0.70 (0.55, 0.89)** | 1.09 (0.72, 1.65) | 1.01 (0.66, 1.54) | 0.89 (0.70, 1.13) | **0.67 (0.48, 0.94)** | 1.09 (0.72, 1.63) |
| cg16885113 | **0.68 (0.52, 0.87)** | 1.09 (0.71, 1.69) | 1.09 (0.70, 1.70) | 0.82 (0.64, 1.05) | **0.66 (0.47, 0.92)** | 1.15 (0.75, 1.76) |
| cg20228636 | **0.70 (0.54, 0.90)** | 1.13 (0.74, 1.74) | 1.11 (0.72, 1.72) | 0.83 (0.65, 1.06) | **0.67 (0.48, 0.94)** | 1.14 (0.75, 1.73) |
| cg25699073 | **0.65 (0.49, 0.86)** | 1.16 (0.69, 1.94) | 1.05 (0.62, 1.79) | **0.74 (0.58, 0.95)** | **0.69 (0.49, 0.96)** | 1.10 (0.67, 1.81) |
| cg25978138 | **0.70 (0.54, 0.91)** | 0.99 (0.62, 1.58) | 1.19 (0.72, 1.96) | **0.74 (0.58, 0.96)** | **0.63 (0.46, 0.88)** | 1.17 (0.73, 1.90) |
| cg13835168 | 0.79 (0.63, 1.00) | 1.24 (0.83, 1.85) | 1.10 (0.75, 1.63) | 0.97 (0.77, 1.23) | **0.70 (0.49, 0.98)** | 1.12 (0.77, 1.63) |
| **AA Participants (Visit 02)** |  |  |  |  |  |  |
| cg07134666 | 1.11 (0.93, 1.31) | 1.00 (0.81, 1.23) | 0.96 (0.81, 1.14) | 1.10 (0.90, 1.33) | 1.02 (0.84, 1.24) | 0.96 (0.79, 1.17) |
| cg11383134 | 1.06 (0.89, 1.25) | 0.98 (0.79, 1.20) | 0.96 (0.81, 1.14) | 1.04 (0.85, 1.26) | 0.96 (0.79, 1.17) | 0.93 (0.76, 1.14) |
| cg22494932 | 1.06 (0.88, 1.26) | 0.95 (0.77, 1.17) | 0.94 (0.79, 1.12) | 1.07 (0.87, 1.30) | 0.96 (0.79, 1.18) | 0.93 (0.76, 1.14) |
| cg15570656 | 1.09 (0.92, 1.29) | 0.97 (0.79, 1.19) | 0.93 (0.78, 1.10) | 1.09 (0.90, 1.32) | 1.00 (0.82, 1.22) | 0.93 (0.76, 1.13) |
| cg16885113 | 1.10 (0.92, 1.30) | 0.98 (0.80, 1.21) | 0.97 (0.82, 1.15) | 1.08 (0.89, 1.32) | 1.00 (0.82, 1.21) | 0.96 (0.79, 1.17) |
| cg20228636 | 1.11 (0.94, 1.32) | 1.00 (0.82, 1.23) | 0.95 (0.81, 1.13) | 1.09 (0.90, 1.32) | 1.02 (0.84, 1.25) | 0.95 (0.78, 1.15) |
| cg25699073 | 1.09 (0.92, 1.30) | 1.01 (0.82, 1.24) | 0.99 (0.83, 1.17) | 1.08 (0.89, 1.31) | 1.01 (0.83, 1.23) | 0.98 (0.80, 1.19) |
| cg25978138 | 1.06 (0.88, 1.26) | 1.01 (0.81, 1.25) | 0.92 (0.77, 1.09) | 1.09 (0.88, 1.33) | 0.99 (0.81, 1.22) | 0.91 (0.75, 1.12) |
| cg13835168 | 1.07 (0.90, 1.27) | 1.00 (0.81, 1.23) | 0.94 (0.79, 1.12) | 1.07 (0.88, 1.30) | 1.02 (0.84, 1.24) | 0.94 (0.77, 1.15) |

<sup>a</sup> A *ZFP57* DMR identified by our DMRcate analysis and also reported in Hernandez et al.
<sup>b</sup> two definitions of PD were used CDC-AAP and ARIC. See Methods.
<sup>c</sup> Model adjusted for age, sex, diabetes, SES, pack-years, smoking status (never, former, current), pack-years based methylation score, BMI, surrogate variables for batch effects, and cell proportions.
NOTE: Bold OR and CI values indicate statistical significance.

**Table 3.** Odds ratio for CpGs in the *ZFP57* ^a^ differentially methylated region and periodontitis severity stratified by age at blood draw at Visit 2, ARIC participants.

| CpG | Odds Ratio per 1 SD increase in methylation level (95% CI) for periodontitis (ARIC definition) |  |  |  |  |  |
| --- | --- | --- | --- | --- | --- | --- |
|  | Age<58<sup>a</sup> |  |  | Age≥58<sup>a</sup> |  |  |
|  | moderate vs. no/mild | severe vs. no/mild | edentulism vs. no/mild | moderate vs. no/mild | severe vs. no/mild | edentulism vs. no/mild |
| **EA Participants (Visit 02)** |  |  |  |  |  |  |
| cg07134666 | 1.01 (0.69, 1.46) | 1.09 (0.60, 1.97) | 0.84 (0.33, 2.10) | **0.59 (0.41, 0.85)** | **0.36 (0.22, 0.60)** | 0.79 (0.48, 1.30) |
| cg11383134 | 0.87 (0.60, 1.26) | 0.93 (0.52, 1.67) | 1.03 (0.37, 2.89) | **0.58 (0.39, 0.86)** | **0.36 (0.22, 0.61)** | 0.82 (0.46, 1.46) |
| cg22494932 | 0.80 (0.56, 1.16) | 1.01 (0.56, 1.82) | 1.10 (0.37, 3.27) | **0.59 (0.40, 0.87)** | **0.40 (0.24, 0.66)** | 0.85 (0.46, 1.60) |
| cg15570656 | 1.00 (0.69, 1.45) | 1.00 (0.56, 1.79) | 0.84 (0.33, 2.12) | **0.69 (0.48, 0.99)** | **0.39 (0.23, 0.65)** | 0.88 (0.52, 1.48) |
| cg16885113 | 0.95 (0.65, 1.37) | 1.04 (0.58, 1.87) | 1.01 (0.38, 2.70) | **0.60 (0.41, 0.88)** | **0.36 (0.21, 0.59)** | 0.83 (0.48, 1.43) |
| cg20228636 | 0.94 (0.64, 1.36) | 0.89 (0.49, 1.60) | 0.90 (0.35, 2.34) | **0.63 (0.44, 0.92)** | **0.42 (0.25, 0.70)** | 0.90 (0.53, 1.53) |
| cg25699073 | 0.75 (0.51, 1.09) | 0.96 (0.53, 1.74) | 1.17 (0.37, 3.70) | **0.64 (0.44, 0.94)** | **0.42 (0.26, 0.70)** | 0.80 (0.44, 1.45) |
| cg25978138 | 0.86 (0.57, 1.30) | 0.84 (0.44, 1.61) | 1.10 (0.36, 3.40) | **0.61 (0.42, 0.89)** | **0.46 (0.30, 0.72)** | 0.97 (0.54, 1.74) |
| cg13835168 | 1.27 (0.87, 1.85) | 1.05 (0.58, 1.91) | 1.01 (0.40, 2.54) | **0.66 (0.46, 0.94)** | **0.42 (0.25, 0.70)** | 0.91 (0.56, 1.49) |
| **AA Participants (Visit 02)** |  |  |  |  |  |  |
| cg07134666 | 1.22 (0.97, 1.55) | 1.18 (0.93, 1.50) | 1.02 (0.80, 1.30) | 0.81 (0.55, 1.21) | 0.68 (0.46, 1.01) | 0.68 (0.46, 1.00) |
| cg11383134 | 1.12 (0.89, 1.41) | 1.10 (0.86, 1.40) | 0.96 (0.75, 1.23) | 0.83 (0.55, 1.25) | **0.65 (0.44, 0.98)** | 0.70 (0.47, 1.04) |
| cg22494932 | 1.09 (0.86, 1.38) | 1.06 (0.83, 1.35) | 0.93 (0.73, 1.19) | 1.00 (0.65, 1.53) | 0.74 (0.49, 1.11) | 0.77 (0.52, 1.15) |
| cg15570656 | 1.18 (0.94, 1.50) | 1.16 (0.91, 1.48) | 0.98 (0.76, 1.26) | 0.85 (0.58, 1.26) | **0.67 (0.45, 0.99)** | **0.68 (0.46, 0.99)** |
| cg16885113 | 1.17 (0.93, 1.48) | 1.13 (0.89, 1.43) | 0.98 (0.77, 1.26) | 0.87 (0.58, 1.29) | 0.70 (0.47, 1.04) | 0.73 (0.50, 1.07) |
| cg20228636 | 1.13 (0.90, 1.43) | 1.15 (0.91, 1.47) | 0.95 (0.74, 1.22) | 0.93 (0.64, 1.36) | 0.73 (0.50, 1.06) | 0.75 (0.52, 1.09) |
| cg25699073 | 1.12 (0.89, 1.41) | 1.13 (0.89, 1.44) | 0.98 (0.77, 1.24) | 0.94 (0.62, 1.42) | 0.74 (0.49, 1.11) | 0.79 (0.53, 1.18) |
| cg25978138 | 1.14 (0.89, 1.45) | 1.05 (0.82, 1.35) | 0.93 (0.73, 1.20) | 0.98 (0.65, 1.47) | 0.82 (0.55, 1.22) | 0.74 (0.50, 1.08) |
| cg13835168 | 1.16 (0.92, 1.47) | 1.19 (0.93, 1.52) | 0.98 (0.76, 1.26) | 0.85 (0.58, 1.23) | **0.67 (0.46, 0.99)** | 0.72 (0.49, 1.03) |

<sup>a</sup> A *ZFP57* DMR identified by our DMRcate analysis and also reported in Hernandez et al.<sup>b</sup> Model adjusted for age, sex, diabetes, SES, pack-years, smoking status (never, former, current), pack-years based methylation score, BMI, surrogate variables for batch effects, and cell proportions.
NOTE: Bold OR and CI values indicate statistical significance.

#### HOXA4

A *HOXA4* DMR, consisting of 13 CpGs, was found to be hypermethylated in periodontitis patients with age-matched and sex-matched periodontally healthy controls.^10^ This finding is consistent with Hernandez et al. While the *HOXA4* DMR was not statistically significant in the DMRcate analyses using both of our periodontitis definitions, when we tested the CpGs contained in the *HOXA4* DMR individually, increasing methylation of cg11015251, cg14359292, cg07317062, and cg08657492 was statistically significantly associated with a higher risk of severe periodontal disease compared to no/mild periodontal disease in African American participants (Table 4). This association was comparable for African American participants 58 years or older and younger than 58-years-old, male and female participants, and participants of higher or lower life-course SES (Supplemental Table 4). In contrast, these *HOXA4* CpGs were not significantly associated with risk of moderate or severe periodontal disease, as compared to no/mild periodontal disease, in European American participants (Table 4). In addition, increasing methylation of cg14359292 and cg24169822 was associated with decreased risk of edentulism vs. no periodontal disease among European American participants (Table 4; stronger association in females Supplemental Table 4). Using the CDC-AAP definition, we also found significant positive associations between increasing methylation of selected *HOXA4* CpGs and periodontitis that were largely consistent with what we found using the ARIC definition.

**Table 4.** Odds ratio for CpGs in the *HOXA4* ^a^ differentially methylated region and periodontitis severity.

| Odds Ratio per 1 SD increase in methylation level (95% CI) |  |  |  |  |  |  |
| --- | --- | --- | --- | --- | --- | --- |
| CpG | Periodontitis (CDC-AAP definition)<sup>b</sup> |  | edentulism vs. no | Periodontitis (ARIC definition)<sup>b</sup> |  |  |
|  | mild/moderate vs. no | severe vs. no |  | moderate vs. no/mild | severe vs. no/mild | edentulism vs. no/mild |
| EA Participants (Visit 02) |  |  |  |  |  |  |
| cg06942814 | 0.92 (0.70, 1.20) | 1.03 (0.66, 1.61) | 0.89 (0.60, 1.34) | 1.03 (0.78, 1.35) | 0.99 (0.66, 1.49) | 0.91 (0.61, 1.35) |
| cg11015251 | 0.83 (0.61, 1.12) | 0.70 (0.43, 1.13) | 0.88 (0.55, 1.39) | 0.97 (0.71, 1.31) | 0.83 (0.55, 1.26) | 0.94 (0.6, 1.45) |
| cg14359292 | 0.85 (0.65, 1.11) | 0.91 (0.60, 1.39) | **0.64 (0.42, 0.99)** | 1.00 (0.77, 1.30) | 1.01 (0.68, 1.50) | 0.70 (0.46, 1.05) |
| cg07317062 | 0.91 (0.71, 1.15) | 1.10 (0.74, 1.64) | 0.93 (0.64, 1.36) | 1.05 (0.82, 1.33) | 1.15 (0.81, 1.65) | 0.99 (0.69, 1.44) |
| cg08657492 | 0.86 (0.66, 1.11) | 0.94 (0.62, 1.42) | 0.70 (0.47, 1.05) | 1.03 (0.79, 1.33) | 0.96 (0.66, 1.42) | 0.75 (0.50, 1.12) |
| cg11410718 | 0.94 (0.74, 1.20) | 1.00 (0.73, 1.66) | 0.86 (0.58, 1.28) | 1.05 (0.82, 1.35) | 1.06 (0.74, 1.51) | 0.89 (0.60, 1.30) |
| cg22997113 | 0.97 (0.75, 1.25) | 1.11 (0.72, 1.71) | 0.91 (0.61, 1.37) | 1.10 (0.85, 1.43) | 1.01 (0.68, 1.49) | 0.92 (0.62, 1.37) |
| cg24169822 | **0.77 (0.59, 1.00)** | 0.83 (0.55, 1.25) | **0.59 (0.39, 0.90)** | 1.03 (0.79, 1.33) | 0.87 (0.59, 1.28) | 0.66 (0.44, 1.01) |
| AA Participants (Visit 02) |  |  |  |  |  |  |
| cg06942814 | **1.20 (1.00, 1.44)** | 1.02 (0.81, 1.27) | 0.94 (0.78, 1.13) | 1.15 (0.93, 1.42) | 1.18 (0.95, 1.46) | 0.98 (0.79, 1.22) |
| cg11015251 | 1.18 (0.97, 1.43) | 0.98 (0.78, 1.24) | 0.93 (0.76, 1.13) | **1.29 (1.03, 1.61)** | **1.28 (1.02, 1.61)** | 1.07 (0.85, 1.34) |
| cg14359292 | 1.18 (0.98, 1.41) | 1.08 (0.87, 1.34) | 0.98 (0.81, 1.17) | 1.17 (0.95, 1.44) | **1.24 (1.01, 1.54)** | 1.05 (0.85, 1.30) |
| cg07317062 | 1.17 (0.98, 1.40) | 1.10 (0.88, 1.37) | 0.98 (0.81, 1.17) | **1.23 (1.00, 1.51)** | **1.30 (1.05, 1.61)** | 1.09 (0.88, 1.34) |
| cg08657492 | **1.25 (1.04, 1.49)** | 1.07 (0.85, 1.33) | 0.99 (0.82, 1.19) | 1.22 (0.99, 1.50) | **1.25 (1.01, 1.55)** | 1.06 (0.86, 1.31) |
| cg11410718 | 1.15 (0.96, 1.38) | 1.10 (0.88, 1.38) | 0.95 (0.79, 1.15) | 1.19 (0.96, 1.46) | 1.20 (0.97, 1.49) | 1.02 (0.82, 1.26) |
| cg22997113 | 1.14 (0.95, 1.36) | 1.04 (0.83, 1.29) | 0.93 (0.78, 1.11) | 1.10 (0.90, 1.35) | 1.13 (0.92, 1.39) | 0.96 (0.78, 1.18) |
| cg24169822 | **1.20 (1.00, 1.44)** | 1.08 (0.86, 1.34) | 0.99 (0.82, 1.19) | 1.19 (0.96, 1.47) | 1.23 (0.99, 1.52) | 1.05 (0.85, 1.31) |

<sup>a</sup> A *HOXA4* DMR reported as periodontitis-related in Hernandez et al.<sup>b</sup> Model adjusted for age, sex, diabetes, SES, pack-years, smoking status (never, former, current), pack-years based methylation score, BMI, surrogate variables for batch effects, and cell proportions.
NOTE: Bold OR and CI values indicate statistical significance.

#### ENSG00000231601

Using the ARIC periodontitis definition, our DMRcate analysis identified one statistically significant DMR located on Chr10 (*ENSG0000023160* gene), consisting of three CpGs (cg22954052, cg25580864, and cg05495790), that differed between African American participants with edentulism and those with no/mild periodontal disease (Table 5). Among African American participants, increasing methylation of cg22954052 and cg05495790 was significantly associated with increased risks of moderate periodontal disease, severe periodontal disease, and edentulism, when compared to no/mild periodontal disease. These associations were stronger among African American participants under age 58 (Supplemental Table 5). cg22954052 and cg05495790 were associated with an increased risk of edentulism vs. no periodontal disease in African American participants. These CpGs were not associated with PD or edentulism in European American participants. When we used the CDC-AAP definition, increasing methylation of cg22954052 and cg05495790 was also significantly associated with increased risks of mild/moderate vs. no periodontal disease and edentulism vs. no periodontal disease, but not severe PD vs no PD, in African American participants.

**Table 5.** Odds ratio for CpGs in the *ENSG00000231601* ^a^ differentially methylated region and periodontitis severity.

| Odds Ratio per 1 SD increase in methylation level (95% CI) |  |  |  |  |  |  |
| --- | --- | --- | --- | --- | --- | --- |
| CpG | Periodontitis (CDC-AAP definition)<sup>b</sup> |  | edentulism vs. no | Periodontitis (ARIC definition)<sup>b</sup> |  |  |
|  | mild/moderate vs. no | severe vs. no |  | moderate vs. no/mild | severe vs. no/mild | edentulism vs. no/mild |
| EA Participants (Visit 02) |  |  |  |  |  |  |
| cg22954052 | 1.11 (0.86, 1.43) | 1.33 (0.87, 2.04) | 0.88 (0.60, 1.29) | 1.08 (0.84, 1.40) | 1.06 (0.74, 1.51) | 0.84 (0.58, 1.23) |
| cg25580864 | 1.10 (0.84, 1.43) | 0.98 (0.64, 1.49) | 1.01 (0.66, 1.53) | 1.03 (0.78, 1.34) | 1.17 (0.77, 1.76) | 1.01 (0.67, 1.52) |
| cg05495790 | 1.12 (0.86, 1.44) | **1.76 (1.16, 2.67)** | 0.85 (0.56, 1.31) | 1.22 (0.94, 1.58) | 1.21 (0.85, 1.72) | 0.84 (0.56, 1.27) |
| AA Participants (Visit 02) |  |  |  |  |  |  |
| cg22954052 | **1.23 (1.02, 1.48)** | 1.18 (0.94, 1.49) | **1.29 (1.07, 1.56)** | **1.54 (1.24, 1.91)** | **1.44 (1.16, 1.79)** | **1.56 (1.25, 1.94)** |
| cg25580864 | 1.17 (0.96, 1.44) | 1.11 (0.87, 1.43) | 1.22 (0.99, 1.50) | 1.16 (0.91, 1.46) | **1.28 (1.01, 1.63)** | **1.30 (1.03, 1.65)** |
| cg05495790 | **1.34 (1.10, 1.63)** | 1.22 (0.95, 1.56) | **1.38 (1.13, 1.69)** | **1.43 (1.13, 1.80)** | **1.36 (1.08, 1.73)** | **1.53 (1.21, 1.94)** |

<sup>a</sup> A de novo *ENSG00000231601* DMR identified by our DMRcate analysis.
<sup>b</sup> Model adjusted for age, sex, diabetes, SES, pack-years, smoking status (never, former, current), pack-years based methylation score, BMI, surrogate variables for batch effects, and cell proportions.
NOTE: Bold OR and CI values indicate statistical significance.

## Discussion

To our knowledge, this is the first large-scale epigenome-wide DNA methylation study of clinically assessed periodontitis. Using epigenome-wide analyses in 1534 ARIC participants, we identified differential methylation patterns comparing various clinically assessed periodontal disease classifications, suggesting that epigenetic changes may contribute to variation in susceptibility to periodontitis. We replicated findings from a previous study reporting an association between DNA methylation levels measured in whole blood at CpG sites in the Zinc Finger Protein Gene 57 (*ZFP57*) and periodontitis using two clinically assessed periodontal disease definitions. We also found DNA methylation differences near *ENSG00000231601* (Chr10) between African American participants with edentulism and those with no or mild periodontal disease.

Our analysis used two periodontitis classifications, ARIC definition and CDC definition, to better handle the complexity of measuring this disease. Since periodontitis is a chronic disease, patients present a wide range of severity of symptoms and progression. Results varied slightly by periodontitis definition used; findings were stronger with the ARIC definition for each of the regions identified. The ARIC definition for “severe” is based on having extensive periodontal disease (30% of teeth with attachment loss ≥3 mm). In contrast, the CDC definition for “severe” requires a greater degree of attachment loss (>6 mm) but fewer teeth. Thus, the different results suggest that the genetic regions may be more strongly linked to extensive periodontal disease. Overall, we found comparable results for male and female participants and participants of higher or lower SES, but found greater associations among participants 58 or older, possibly due to a higher prevalence of extensive periodontal disease among older ARIC participants. The differences we observed between African Americans and European Americans may have been due to differences in the two populations. The large majority of African American participants are from the Jackson Field Center. Participants from this region experienced very different exposures during their lifetimes and have limited access to health and dental care (compared to other ARIC participants). These factors, which can influence periodontal disease development and progression, may have impacted findings in a manner similar to differences we observed with different disease classifications. Consequently, it is not possible to determine whether our results are due to genetic effects or correlates of socioeconomic factors.

Prior to the present study, there were only two published studies of genome-wide DNA methylation of peripheral blood leukocytes in periodontitis. Kurushima et al. (2019) conducted an epigenome-wide study using Illumina DNA methylation 450K data and a retrospective dental phenotype collection of self-reported dental mobility and gingival bleeding information obtained from 528 older female twins in the Twins UK cohort (mean age: 58 years old, age range: 19-82 years old; compared to African American ARIC participants mean age: 55 years old, age range: 47-70 years old, European American ARIC participants mean age: 58 years old, age range: 47-68 years old).^9^ This study detected a hypomethylated region in *ZNF804A* in individuals who experience gingival bleeding, in conjunction with an increased level of ornithine, a metabolite related to gingival inflammation, in blood metabolomics profile. There were no concordances between our results and the results of Kurushima et al., possibly due to clear differences in study design, including the use of clinically examined vs. self-reported periodontitis data. Hernandez et al. (2021) performed an analysis to identify differentially methylated positions and regions with a small number of clinically diagnosed periodontitis patients and gingivally healthy controls^10^. Results of Hernandez et al. identified *ZNF718, HOXA4*, and *ZFP57* genes related to systemic immune-related epigenetic patterns in periodontitis^10^, including a *ZFP57* region of 21 CpGs that was found to be hypomethylated when comparing periodontitis patients with age-matched and sex-matched periodontally healthy controls. While Hernandez et al. also carried out clinical examination, its study participants were much younger than ours (age range: 25-55 years old), suggesting a more extreme comparison of phenotype in Hernandez et al. Consistent with Hernandez et al., our study identified a *ZFP57* DMR to be periodontitis-related. In addition, the 17 CpGs of this selected DMR perfectly overlapped with the 21-CpG *ZFP57* DMR identified by Hernandez and colleagues.

Existing evidence supports the relevance of some of the identified *ZFP57* CpGs for periodontitis. *ZFP57* is an imprint regulator, and DNA methylation in this exact *ZFP57* region has been shown to be environmentally modulated in human blood and related to transient neonatal diabetes, Parkinson’s disease, and post-traumatic stress disorder.^30-32^ The Zinc Finger Proteins (ZNFs) are also important transcription factors. They are involved in tissue development, and their alterations may promote chronic conditions, including oral cancer and periodontitis.^33^ Recent studies have shown ZNFs playing a role in the progression of periodontitis.^34^ For instance, *ZNFA20* can inhibit NF-κβ pathway activation via inflammatory cytokines^34,35^ and thus support the disruption of local inflammatory responses by *P. gingivalis*, the keystone pathogen of periodontitis.^36,37^ The replication of the *ZNF57* DMR in our study suggests a potential sequence of events in periodontitis development, where hypomethylation of the *ZNF57* DMR may upregulate the expression of *ZNF57*, which is related to antigen presentation and immune response regulation.

Observed DNA methylation levels could precede periodontal disease development or be the consequence of periodontal disease. The main limitation of the present study is that we could not establish whether associations we detected between differential methylation in whole blood and periodontitis were the cause rather than the consequences of periodontitis. While ARIC is a prospective study and the methylation data were from blood samples collected up to six years before ARIC dental examination, a dental examination was not performed concurrently with the blood draw, such that we likely studied the combination of prevalent and incident periodontal disease. Moreover, we could not detect single CpG associations at the stringent epigenome-wide significance level. Another limitation is using the 450K array, which covers only ∼2% of human CpG sites and limits the discovery. Lastly, our use of edentulism as a surrogate for the most severe type of periodontitis is not necessarily accurate, given that the cause of edentulism is multi-faceted, including but not limited to having and being treated for periodontal disease and poor access to dental care. Consequently, our analysis cannot differentiate DNA methylation patterns of those who are edentulous due to having periodontitis vs. those who are edentulous for other reasons.

Despite the limitations, several strengths of this study warrant attention, including the use of clinically examined periodontal phenotypes, large sample size, ethnic diversity, and rigorous analytical methods. We also adjusted for SES, diabetes status, imputed cell composition, self-reporting smoking history, and a methylation-predicted packyears smoked to further reduce concern about confounding. Additionally, we utilized two definitions of periodontal disease to address the complexity of disease classification. Analyses in individuals of different ancestries support the need for future large-scale EWAS of periodontitis to investigate ethnicity-specific and age-specific periodontitis-related CpGs.

Along with previously published EWAS, our results suggest that gene regulation mechanisms may influence the susceptibility to and severity of periodontitis. Our well-powered EWAS and DMRcate analysis in participants drawn from the general population identified several genomic regions for which differential DNA methylation sites were associated with periodontitis. This study supports leukocyte DNA methylation for the evaluation of epigenetic patterns in periodontitis. Our study, in conjunction with a prior pilot study, implicates *ZFP57* as a promising candidate for future studies to illuminate the underlying gene regulatory mechanisms linking differential DNA methylation to periodontal disease.

## Data Availability

The datasets used in the current study are available from the ARIC study.

## Reference

1. Eke P, Dye B, Wei L, Thornton-Evans G, Genco R. CDC Periodontal Disease Surveillance workgroup: James Beck (University of North Carolina, Chapel Hill, USA). Gordon Douglass (Past President, American Academy of Periodontology), Roy Page (University of Washin Prevalence of periodontitis in adults in the United States: 2009 and 2010 J Dent Res. 2012;91(10):914–920.

2. Eke PI, Dye BA, Wei L, et al. Update on prevalence of periodontitis in adults in the United States: NHANES 2009 to 2012. Journal of periodontology. 2015;86(5):611–622.

3. Eke PI, Borgnakke WS, Genco RJ. Recent epidemiologic trends in periodontitis in the USA. Periodontology 2000. 2020;82(1):257–267.

4. Slots J. Periodontitis: facts, fallacies and the future. Periodontology 2000. 2017;75(1):7–23.

5. Petersen PE, Ogawa H. The global burden of periodontal disease: towards integration with chronic disease prevention and control. Periodontology 2000. 2012;60(1):15–39.

6. Laine ML, Jepsen S, Loos BG. Progress in the identification of genetic factors in periodontitis. Current Oral Health Reports. 2014;1(4):272–278.

7. Divaris K, Monda KL, North KE, et al. Exploring the genetic basis of chronic periodontitis: a genome-wide association study. Human molecular genetics. 2013;22(11):2312–2324.

8. Hajishengallis G. Periodontitis: from microbial immune subversion to systemic inflammation. Nature reviews immunology. 2015;15(1):30–44.

9. Kurushima Y, Tsai P-C, Castillo-Fernandez J, et al. Epigenetic findings in periodontitis in UK twins: a cross-sectional study. Clinical epigenetics. 2019;11(1):1–13.

10. Hernández HG, HernándezLJCastañeda AA, Pieschacón MP, Arboleda H. ZNF718, HOXA4, and ZFP57 are differentially methylated in periodontitis in comparison with periodontal health: epigenomeLJwide DNA methylation pilot study. Journal of Periodontal Research. 2021;56(4):710–725.

11. Wright JD, Folsom AR, Coresh J, et al. The ARIC (atherosclerosis risk in communities) study: JACC focus seminar 3/8. Journal of the American College of Cardiology. 2021;77(23):2939–2959.

12. Jackson R, Chambless LE, Yang K, et al. Differences between respondents and nonrespondents in a multicenter community-based study vary by gender and ethnicity. Journal of clinical epidemiology. 1996;49(12):1441–1446.

13. Naorungroj S, Slade G, Divaris K, Heiss G, Offenbacher S, Beck J. Racial differences in periodontal disease and 10LJyear selfLJreported tooth loss among late middleLJaged and older adults: the dental Aric study. Journal of public health dentistry. 2017;77(4):372–382.

14. Bose M, Wu C, Pankow JS, et al. Evaluation of microarray-based DNA methylation measurement using technical replicates: the Atherosclerosis Risk In Communities (ARIC) Study. BMC bioinformatics. 2014;15(1):1–10.

15. Michaud DS, Lu J, Peacock-Villada AY, et al. Periodontal disease assessed using clinical dental measurements and cancer risk in the ARIC study. JNCI: Journal of the National Cancer Institute. 2018;110(8):843–854.

16. Beck JD, Eke P, Heiss G, et al. Periodontal disease and coronary heart disease: a reappraisal of the exposure. Circulation. 2005;112(1):19–24.

17. Beck JD, Elter JR, Heiss G, Couper D, Mauriello SM, Offenbacher S. Relationship of periodontal disease to carotid artery intima-media wall thickness: the atherosclerosis risk in communities (ARIC) study. Arteriosclerosis, thrombosis, and vascular biology. 2001;21(11):1816–1822.

18. Eke PI, Page RC, Wei L, ThorntonLJEvans G, Genco RJ. Update of the case definitions for populationLJbased surveillance of periodontitis. Journal of periodontology. 2012;83(12):1449–1454.

19. Lazăr L, Dakó T, Cozma A, Lazăr A-P. The influence of smoking on the periodontal biome. A review. Acta Stomatologica Marisiensis Journal.5(1):6–11.

20. Martinon P, Fraticelli L, Giboreau A, Dussart C, Bourgeois D, Carrouel F. Nutrition as a key modifiable factor for periodontitis and main chronic diseases. Journal of Clinical Medicine. 2021;10(2):197.

21. Spector AM, Postolache TT, Akram F, Scott AJ, Wadhawan A, Reynolds MA. Psychological stress: a predisposing and exacerbating factor in periodontitis. Current Oral Health Reports. 2020;7(3):208–215.

22. Genco RJ, Borgnakke WS. Diabetes as a potential risk for periodontitis: association studies. Periodontology 2000. 2020;83(1):40–45.

23. Madiba TK, Bhayat A. Periodontal disease-risk factors and treatment options. South African Dental Journal. 2018;73(9):571–575.

24. Lu J, Zaimi I, Barber JR, et al. SES and correlated factors do not explain the association between periodontal disease, edentulism, and cancer risk. Ann Epidemiol. 2019;38:35–41.

25. Plerou V, Gopikrishnan P, Rosenow B, Amaral LAN, Guhr T, Stanley HE. Random matrix approach to cross correlations in financial data. Physical Review E. 2002;65(6):066126.

26. Leek JT, Scharpf RB, Bravo HC, et al. Tackling the widespread and critical impact of batch effects in high-throughput data. Nature Reviews Genetics. 2010;11(10):733–739.

27. Joehanes R, Just AC, Marioni RE, et al. Epigenetic Signatures of Cigarette Smoking. Circ Cardiovasc Genet. 2016;9(5):436–447.

28. Peters TJ, Buckley MJ, Statham AL, et al. De novo identification of differentially methylated regions in the human genome. Epigenetics & chromatin. 2015;8(1):1–16.

29. Benjamini Y, Hochberg Y. Controlling the false discovery rate: a practical and powerful approach to multiple testing. Journal of the Royal statistical society: series B (Methodological). 1995;57(1):289–300.

30. Bak M, Boonen SE, Dahl C, et al. Genome-wide DNA methylation analysis of transient neonatal diabetes type 1 patients with mutations in ZFP57. BMC medical genetics. 2016;17(1):1–8.

31. Henderson AR, Wang Q, Meechoovet B, et al. DNA methylation and expression profiles of whole blood in parkinson’s disease. Frontiers in Genetics. 2021;12:640266.

32. Vinkers CH, Geuze E, van Rooij SJ, et al. Successful treatment of post-traumatic stress disorder reverses DNA methylation marks. Molecular psychiatry. 2021;26(4):1264–1271.

33. Agnihotri R, Gaur S. The Role of Zinc Finger Proteins in Various Oral Conditions. The Scientific World Journal. 2022;2022.

34. Hong JY, Bae WJ, Yi JK, Kim GT, Kim EC. AntiLJinflammatory and antiLJosteoclastogenic effects of zinc finger protein A 20 overexpression in human periodontal ligament cells. Journal of periodontal research. 2016;51(4):529–539.

35. Wertz IE, O’rourke KM, Zhou H, et al. De-ubiquitination and ubiquitin ligase domains of A20 downregulate NF-κB signalling. Nature. 2004;430(7000):694–699.

36. Ohshima J, Wang Q, Fitzsimonds ZR, et al. Streptococcus gordonii programs epithelial cells to resist ZEB2 induction by Porphyromonas gingivalis. Proceedings of the National Academy of Sciences. 2019;116(17):8544–8553.

37. Matsui S, Zhou L, Nakayama Y, et al. MiR-200b attenuates IL-6 production through IKKβ and ZEB1 in human gingival fibroblasts. Inflammation Research. 2018;67(11):965–973.

