## Supplemental Methods for "Epigenome-wide association study using peripheral blood leukocytes identifies genomic regions associated with periodontal disease and edentulism in the Atherosclerosis Risk in Communities Study"

**Supplementary Methods**

**DNA Methylation Assessing and Preprocessing**

Genomic DNA was extracted from peripheral blood leukocyte samples using the Gentra Puregene Blood Kit (Qiagen) while bisulphite conversion of 1 ug genomic DNA was performed using the EZ-96 DNA Methylation Kit (Deep Well Format) (Zymo Research). Genome-wide profiling of DNA methylation was assayed using the Illumina Infinium HumanMethylation450K BeadChip array (HM450K) in 2,853 African American participants at 483,525 CpG sites and in 1,104 European American participants at 482,815 CpG sites. The degree of methylation, background subtraction, and average normalization was determined and conducted using Illumina GenomeStudio 2011.1, Methylation module 1.9.0 software.

**Estimation of peripheral blood leukocyte composition**

Peripheral blood leukocyte subtypes proportions, including myeloid lineage sub-types [neutrophils (Neu), eosinophils (Eos), basophils (Bas), and monocytes (Mono)] and lymphoid lineage subtypes [B lymphocytes naïve (Bnv), B lymphocytes memory (Bmem), T helper lymphocytes naïve (CD4nv), T helper lymphocytes memory (CD4mem), T regulatory cells (Treg), T cytotoxic lymphocytes naïve (CD8nv), T cytotoxic lymphocytes memory (CD8mem), and natural killer lymphocytes (NK)], were estimated using a newly expanded reference-based deconvolution library EPIC IDOL-Ext (1). This library used the IDOL methodology (2) to optimize the currently available six-cell reference library (3) to deconvolve the proportions of 12 leukocyte subtypes in peripheral blood. This EPIC IDOL-Ext library (Bioconductor package FlowSorted.BloodExtended.EPIC) was validated using flow cytometry gold standard data and substantiated by including publicly available data from >100,000 samples (1).

**Periodontal Measures**

At visit 4, ARIC participants were asked “do you have any of your natural teeth" and “do you have any dental implants”. If the answer was yes to either, participants who otherwise met the eligibility criteria were examined. If no to both, then the participants were not invited to the dental examination and were classified as edentulous.

During examination, to adjust for AL not due to periodontitis, a correction was made for buccal sites exhibiting >3 mm of AL such that they could not be more than the other sites (adjacent mesio-buccal and disto-buccal) on the same tooth. Dental examiners were calibrated against a standard examiner and each other. The agreement between examiners was very high, as previously reported (4).

For periodontitis CDC-AAP definition, third molars are excluded and CAL and PD measures at four interproximal sites per tooth are included. Periodontal disease severity is classified as follows: No, no evidence of mild, moderate, or severe periodontitis; Mild, >2 interproximal sites with AL>3mm, and >2 interproximal sites with PD>4mm (not on same tooth) or one site with PD>5mm; Moderate, >2 interproximal sites with AL>4mm (not on same tooth), or >2 interproximal sites with PD>5mm (not on same tooth); and Severe, >2 interproximal sites with AL>6mm (not on same tooth) and >1 interproximal site with PD>5mm. For periodontitis ARIC definition (a definition used in a previous study on periodontitis in the ARIC study (5)), periodontitis was categorized using CAL measurements as follows: No/mild periodontitis, <10% of examined sites having AL>3 mm; Moderate periodontitis, >10% to <30% of examined sites having AL>3 mm, and Severe periodontitis, >30% of examined sites with AL>3 mm.

**Pack-years methylation score**

The pack-years methylation score is calculated to represent packyears smoked-associated methylation alterations (6). This packyears methylation score was first developed to predict smoking pack-years using smoking ‘signatures’ reported by large-scale epigenome-wide association meta-analyses (6). This score correlates with gene expression changes that are affected by smoking and can be utilized in lieu of self-reported smoking data (6).

Reference:

1.	Salas LA, Zhang Z, Koestler DC, Butler RA, Hansen HM, Molinaro AM, et al. Enhanced cell deconvolution of peripheral blood using DNA methylation for high-resolution immune profiling. bioRxiv. 2021.

2.	Koestler DC, Jones MJ, Usset J, Christensen BC, Butler RA, Kobor MS, et al. Improving cell mixture deconvolution by identifying optimal DNA methylation libraries (IDOL). BMC bioinformatics. 2016;17(1):120.

3.	Salas LA, Koestler DC, Butler RA, Hansen HM, Wiencke JK, Kelsey KT, et al. An optimized library for reference-based deconvolution of whole-blood biospecimens assayed using the Illumina HumanMethylationEPIC BeadArray. Genome biology. 2018;19(1):1-14.

4.	Beck JD, Eke P, Heiss G, et al. Periodontal disease and coronary heart disease: a reappraisal of the exposure. Circulation 2005;112(1):19-24.

5.	Beck JD, Elter JR, Heiss G, et al. Relationship of periodontal disease to carotid artery intima-media wall thickness: the atherosclerosis risk in communities (ARIC) study. Arterioscler Thromb Vasc Biol 2001;21(11):1816-22.

6.	Sugden K, Hannon EJ, Arseneault L*, et al.* Establishing a generalized polyepigenetic biomarker for tobacco smoking. Translational psychiatry 2019;9(1):1-12.
